# Combining clinical and diagnostic surveillance to estimate the burden of measles disease: a modeling study

**DOI:** 10.1101/2024.05.20.24307625

**Authors:** Tiffany Leung, Matthew Ferrari

**Affiliations:** Center for Infectious Disease Dynamics, Pennsylvania State University, University Park, PA, USA

## Abstract

**Background:** The clinical case definition for measles is highly sensitive and has low specificity. Diagnostic confirmation can resolve this uncertainty for individual cases and is a crucial tool for confirmation of measles outbreaks. However, in under-resourced settings, it is prohibitive to confirm all suspected cases and routine measles surveillance comprises a combination of both clinically and diagnostically confirmed cases.

**Methods:** We developed a dynamic model of measles, rubella, and other sources of febrile rash to simulate time series of a suspected measles surveillance system. We simulated partial reporting of suspected cases and limited routine diagnostic testing using assays with sensitivity and specificity that correspond to current or proposed rapid diagnostic tests. We estimated the time series of reported measles cases as the product of suspected cases and the proportion of diagnostic positive cases. We then estimated the reporting rate and annual incidence for measles using the time-series SIR model.

**Results:** Reconstructing the time series of reported measles cases using the fraction of diagnostic positive cases results in unbiased estimates of the reporting rate and the annual incidence at moderate vaccination levels for all reasonable levels of test sensitivity and specificity, even for low proportions tested. At high vaccination levels, diagnostic tests with low sensitivity (*<* 90%) lead to slight bias in annual incidence. Temporal variation in the prevalence of measles among suspected cases require that the proportion of cases attributable to measles be estimated frequently (i.e., monthly) to avoid bias in estimates.

**Conclusion:** Combining routine, systematic diagnostic confirmation of suspected measles cases with suspected cases surveillance can improve estimates of the reporting rate and annual incidence using diagnostic tests with sensitivity and specificity consistent with proposed rapid diagnostic tests.

## 1 Introduction

Measles remains a major burden in many countries despite the use of measles vaccine since the late 1960s [1]. The measles clinical case definition is a general maculopapular rash lasting at least 3 days, fever, and at least one of the following: cough, coryza, or conjunctivitis [2]. When measles incidence is high, either in settings with low vaccination or during times of outbreaks, a large fraction of febrile-rash cases is attributable to measles and the positive predictive value of the clinical case definition is comparatively high. When measles incidence is low, clinical surveillance becomes dominated by other causes of febrile rash, such as rubella infection, and may become an unreliable indicator for the absolute burden of disease [2].

Measles infection can be diagnostically confirmed using a capture IgM enzyme immunoassay (EIA), viral culture, or RT-PCR [3]. However, in resource-limited settings, these assays are rarely done on all suspected cases [4]. For example, in the World Health Organization (WHO) African region the annual proportion of suspected measles cases from which samples were collected ranged from 19–81% and the proportion of samples that were IgM-positive for measles ranged from 11–31% [4]. Thus, measles surveillance comprises a combination of records with different diagnostic certainty.

Limited diagnostic confirmation of suspected measles cases can be used to estimate the proportion of untested cases that would be expected to be confirmed, if tested [5]. Bansal et al. [6] and Simonsen et al. [7] have previously argued for the combination of low-specificity data sources (e.g., clinical confirmation and internet searches) with higher specificity measures (e.g., diagnostic confirmation) to generate “hybrid” surveillance that leverages the low cost of the former with the higher specificity of the latter. The minimally sufficient level of high-specificity confirmation to adequately correct for diagnostic uncertainty in low-specificity data sources is likely to be highly system-specific. It would depend on the spatial and temporal variability in the positive predictive value of the low-specificity data source, the sensitivity and specificity of the high-specificity data source, and the particular application (e.g., burden estimation or outbreak detection [8]).

For the purposes of estimating the total burden of measles infection, the clinical case definition for a suspected measles case has overall low specificity that decreases as the relative abundance of measles declines [2]. As measles vaccination increases and prevalence declines, we expect the positive predictive value of the clinical case definition to decline. This will be further impacted by the coverage of rubella-containing vaccine that will impact the prevalence of non-measles sources of febrile rash. Additionally, as measles vaccination increases, the inter-annual variability in measles is expected to increase [9]. Thus, both the mean relative proportion of febrile-rash cases attributable to measles and its variance are expected to change in response to vaccination program improvements.

Rapid diagnostic tests (RDTs) present an opportunity to expand diagnostic confirmation both by reducing the cost per test and removing the cost associated with sample transport to centralized laboratory facilities [10]. Concerns about reduced sensitivity and specificity for RDTs relative to conventional laboratory-based EIAs have limited their adoption for both clinical diagnosis and surveillance [8]. However, recent studies indicate that prototype IgM RDTs can achieve sensitivity and specificity comparable to standard laboratory-based IgM EIA using serum, capillary blood, or oral fluid [8, 11, 12].

In this paper, we evaluate the application of the standard time-series Susceptible-Infected-Recovered (TSIR) model estimator of the burden of measles disease using time series of febrile rash incidence generated by a dynamic multi-pathogen model. We assume that a fraction of reported febrile rash cases is tested for confirmation as measles-positive using a test with sensitivity and specificity consistent with the proposed measles IgM RDT target product profile (TPP) and the remainder are recorded as clinically compatible, suspected measles cases. We then quantify the performance of the estimators of the reporting rate and total burden of measles infection.

## 2 Methods

### 2.1 Model of measles and rubella transmission with vaccination

We constructed an age-structured stochastic compartmental model of measles and rubella transmission with measles vaccination. The population is divided into 40 age groups (24 monthly groups between 0 and 2 years old; 1 group for 2 to 5 years old; 14 five-year groups thereafter until 75 years old, and 1 group for 75+ years). Individuals are either unvaccinated or vaccinated, and within each vaccination status, are further classified by immune status each to measles and to rubella as immune with maternal antibodies (M), susceptible (S), infectious (I), or recovered (R).

Individuals are born either susceptible or temporarily immune with maternal antibodies; the fraction of the birth cohort at time *t* that is born with maternal antibodies is proportional to the proportion of individuals between 10 to 55 years old that are immune, either through prior infection or vaccination [13]. We used a birth rate of 27 births per 1,000 persons per year, consistent with a Kenya-like setting (in the supplement, we illustrate results for simulations with a higher birth rate) [14]. Individuals face a constant death rate, set equal to the birth rate for a constant population size. We modeled aging with discrete age classes and assumed exponential rates of aging occurring every 15 days.

Susceptible individuals can be infected with either measles or rubella. We use an average infectious period of two weeks for both infections. We assume lifelong immunity to measles following measles infection or measles vaccination, and lifelong immunity to rubella following rubella infection. We assume no cross-protection between measles and rubella and disallow coinfection. Thus, while an individual is infected with either measles or rubella, they cannot be infected with the other. We assume either homogeneous contact mixing or assortative contact mixing following the pattern for Kenya in [15]. We assume that the basic reproduction number is 15 for measles [16, 17] and 5.87 for rubella [18]. To re-establish transmission in the event of stochastic fade-out of either measles or rubella, we allow an introduction rate of both in all time steps with probability 0.003.

We model routine vaccination of measles as a one-dose vaccine that is 94% effective against measles infection (and ineffective against rubella) [19]. We use an age-dependent rate of routine vaccination, shaped as a truncated normal distribution with mean at 9 months of age and standard deviation of 0.5 months, constrained to ages 0 and 24 months. The height of the normal distribution is scaled by a constant factor such that the cumulative probability of vaccination by 24 months of age is equal to a specified vaccination coverage. We consider routine vaccination coverage ranging 20% to 90%. If a maternally immune or recovered individual is vaccinated they remain maternally immune or recovered respectively. If a susceptible individual is vaccinated, they become immunized against measles infection with probability 0.94 [19]. Model equations are presented in the Supplementary Information.

We initiate the model with a population size of 1 million with an age structure similar to Kenya [20]. The initial population was fully susceptible to measles and rubella, with 5 individuals infected with measles and susceptible to rubella and 5 individuals infected with rubella and susceptible to measles. We simulate individual time series of length 200 years with vaccination coverage, C, following a discarded transient period of 500 years (300 years without vaccination and 200 years with vaccination at coverage C) to reach equilibrium. We then divide the full time series into 20 individual time series, each 10 years in length.

We use the tau-leaping algorithm [21] and a daily time-step to simulate the system. The number of events is drawn from a Poisson distribution with mean *r*_*i*_, where *r*_*i*_ is the rate of event *i* per day. If the number of events is higher than what is feasible, the maximum allowable number is used.

### 2.2 Estimating the annual incidence of measles

We generate a time series of febrile rash infections due to three sources: measles, rubella, and other non-measles and non-rubella febrile rash. Age distributions of measles and rubella infections are generated from the dynamical model. We add an additional 20% of total infections as febrile rash from other sources, assumed to be geometrically distributed with a mean age of 5 years old. For each 10-year time series, we model imperfect testing and estimate the true burden of measles.

We assume that 10% of all febrile rash infections, regardless of cause, seek care. This reflects a high-performing surveillance system in an endemic region; the estimated proportion of measles cases that are reported in the WHO African, Eastern Mediterranean, South-East Asian, Western Pacific regions in 2022 range from 1–2% [1]. Of the individuals that seek care, we assume a fraction of these cases is selected uniformly at random for diagnostic confirmation with a diagnostic assay that is 90% sensitive and 95% specific, which aligns with the accuracy of current measles IgM rapid diagnostic tests [22]. Those that are not tested are recorded as suspected measles cases. We also simulate a perfect diagnostic test (100% sensitivity and specificity) and tests with lower sensitivity and specificity.

For each simulated time series of febrile rash we estimate the true measles burden in multiple steps. First, we estimate the relative proportion of febrile-rash cases attributable to measles as the fraction of diagnostic tests that are measles-positive. For each simulated day, we estimate the fraction of febrile-rash cases attributable to measles as 1) the fraction positive over the full 10-year time series, 2) the fraction positive over the previous 365 days (annual moving window), and 3) the fraction positive over the previous month (monthly moving window). In addition we assume a counterfactual scenario without testing in which all (suspected measles) febrile-rash cases are treated as true measles cases.

On each day of the time series we take a binomial random draw from those untested suspected untested cases with probability equal to the fraction of febrile-rash cases attributed to measles on each day using each of the methods above. We then generate a reconstructed time series of inferred measles cases that is the sum of the tested cases that were positive and the binomial draw from the untested cases. For each time series, we repeat this 20 times to generate replicate time series that reflect the uncertainty in the binomial reconstruction.

For each reconstructed time series, we estimate the reporting rate following the method described by Finkenstädt and Grenfell [23] as modified by Leung and Ferrari [24]. Briefly, the modified method differs from Finkenstädt and Grenfell [23] by correcting for a bias due to the double counting of cases in individuals too young to be vaccinated. Finally, the unobserved measles burden is calculated by dividing the reconstructed incidence by the reporting rate.

We compare the estimated reporting rates for time series over different vaccination coverage and diagnostic testing rates from 20% to 100%. We assess the performance of the estimates of total measles burden by regressing the estimated annual burden of measles against the true simulated measles burden. A slope of 1 indicates no bias in the estimate of annual burden and an *R*^2^ close to 1 indicates low inter-annual variability in estimates. Though we fit a constant reporting rate, the variability of *R*^2^ reflects variability due to the reconstruction of measles-positive cases from among suspected measles cases due to temporal variability in the relative proportion attributable to measles. For each combination of demographic rates and vaccination coverage, we generate 20 replicate 10-year time series and report the mean and range of estimates.

We also assess the impact of diagnostic test sensitivity and specificity on the estimate of total measles burden under different vaccination coverage. For this part of the analysis, we used a monthly moving window to estimate the fraction of cases attributable to measles. We varied sensitivity and specificity between 85% to 100% and testing rates between 20% to 60%.

## 3 Results

We generated “true” time series of measles, rubella, and other non-measles, non-rubella febrile rash under routine vaccination coverage ranging 20% to 90%. This corresponds to an overall proportion of measles among all febrile-rash cases of 15% (high coverage) to 41% (low coverage) (Table 1). The relative proportion of febrile rash attributable to measles in any given time is variable, even when vaccination is low, because of the occurrence of outbreaks (Figure 1). Here we present results assuming homogeneous contact mixing as that is consistent with the implicit assumptions of the TSIR model, which is not age-specific. The performance of estimators under an age-specific Kenya-like contact mixing is presented in the supplement.

**Table 1:** Percentage of measles among tested cases by test sensitivity-specificity (rows) and vaccination coverage (columns) with 20% testing rate for the full 200 years.

| Sensitivity-Specificity | Vaccination Coverage |  |  |  |  |
| --- | --- | --- | --- | --- | --- |
|  | 20% | 40% | 60% | 80% | 90% |
| True proportion | 0.41 | 0.36 | 0.3 | 0.22 | 0.15 |
| 100-100 | 0.41 | 0.36 | 0.3 | 0.22 | 0.15 |
| 90-95 | 0.39 | 0.35 | 0.3 | 0.23 | 0.18 |
| 90-90 | 0.42 | 0.39 | 0.34 | 0.27 | 0.22 |
| 85-90 | 0.41 | 0.37 | 0.33 | 0.26 | 0.21 |
| 90-85 | 0.46 | 0.42 | 0.38 | 0.31 | 0.26 |
| 85-85 | 0.43 | 0.40 | 0.36 | 0.3 | 0.26 |

**Figure 1.**
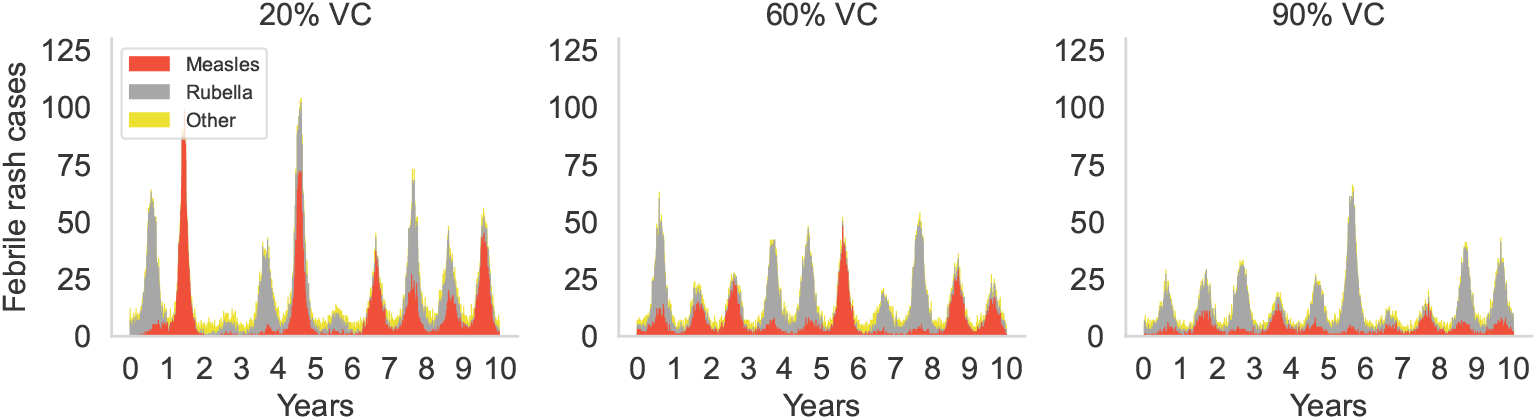
Model-generated clinical febrile rash cases due to measles, rubella, and other non-measles, non-rubella sources over different vaccination coverage (stacked).

As expected, treating all suspected measles cases as true measles cases (scenario without testing) in the TSIR model results in significant bias in the estimate of the reporting rate (Table 2). For a true reporting rate of 10%, this bias increases with vaccination coverage, estimating 23.8% to 77.9% at low and high coverage respectively (Table 2). These estimated reporting rates are used to reconstruct the time series of the true measles infections (Figure 2A–C). Treating all suspected measles cases as true measles cases led to a reconstructed time series that may overestimate measles infections during periods of high rubella infections (e.g., at 20% vaccination coverage, at year 0 and year 3 in Figure 2A and Figure 1). The correlation between true annual measles incidence and estimated annual measles incidence is positive for all levels of vaccination coverage; however, the bias in this relationship (slope of a regression of estimated annual incidence against true annual incidence) increases as vaccination coverage increases and measles becomes a smaller fraction of all observed febrile rash (Table 2; Figure 2 A–C). In addition to the increase in bias at higher vaccination, the inter-annual variability in predictions increases; the coefficient of determination *R*^2^ falls from 0.54 to 0.16 as vaccination increases from 20–90% (Table 3).

**Table 2:**
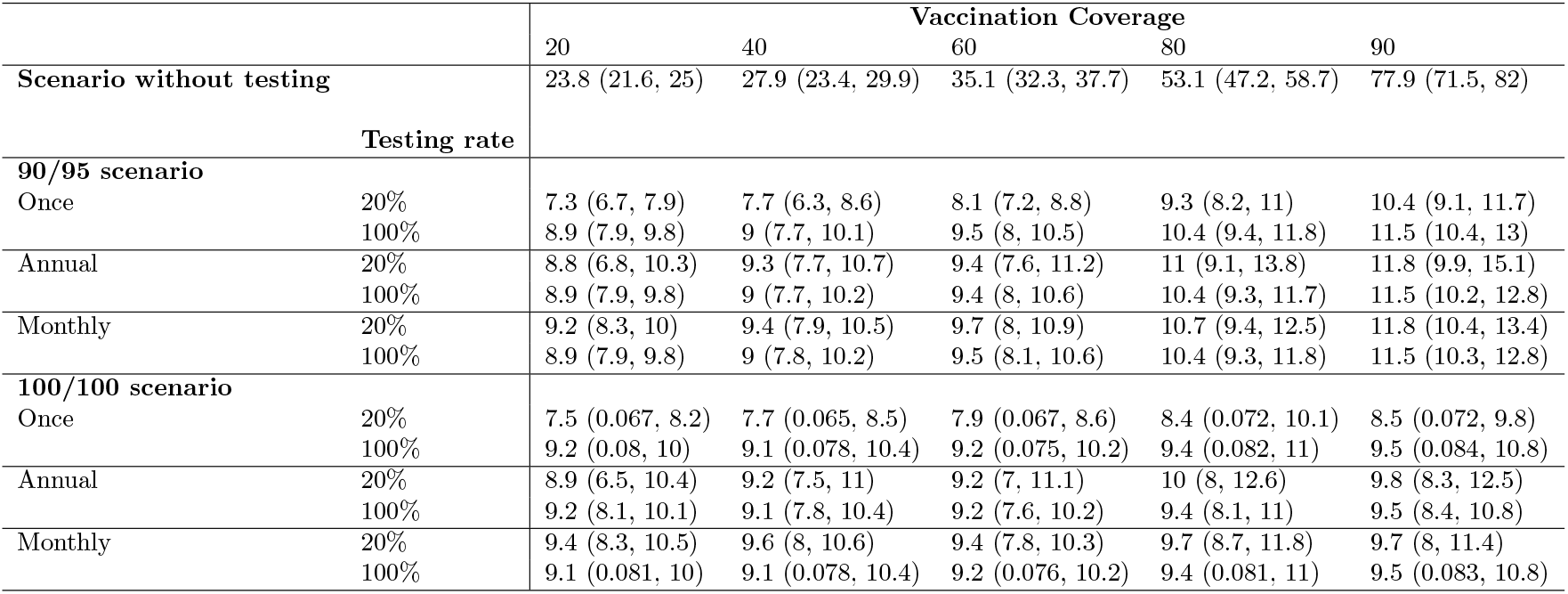
Estimated reporting rates under different measles vaccination coverage and time frequency for estimating the proportion of cases attributable to measles. We considered scenarios without testing, using a test with 90% sensitivity and 95% specificity, and using a test with 100% sensitivity and specificity. The true reporting rate is 10%. Values in brackets show the range over 20 replicate time series.

**Table 3:**
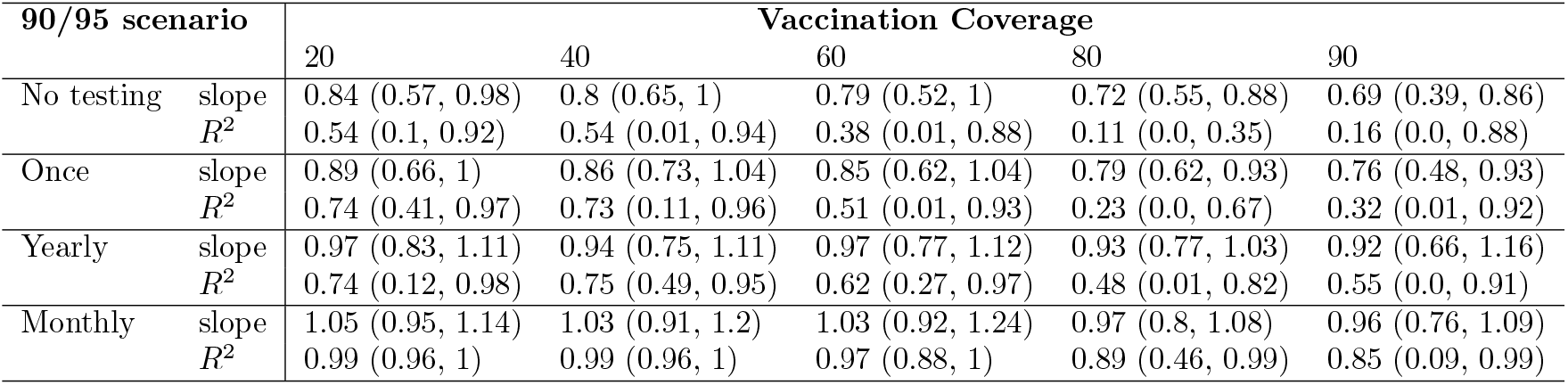
Slope and coefficient of determination (*R*^2^) of the regression of estimated annual incidence of measles against the true simulated incidence of measles infection as a function of measles vaccination coverage (columns) and the time scale for estimating the proportion of cases attributable to measles (rows). Slope near 1 indicates low bias in estimates of annual incidence. *R*^2^ near 1 indicates low variation in estimates from the true values. Values in brackets show the range over 20 replicate time series. (90/95 scenario with 20% sampling)

| 90/95 scenario |  | Vaccination Coverage |  |  |  |  |
| --- | --- | --- | --- | --- | --- | --- |
|  |  | 20 | 40 | 60 | 80 | 90 |
| No testing | slope | 0.84 (0.57, 0.98) | 0.8 (0.65, 1) | 0.79 (0.52, 1) | 0.72 (0.55, 0.88) | 0.69 (0.39, 0.86) |
| | $R^2$ | 0.54 (0.1, 0.92) | 0.54 (0.01, 0.94) | 0.38 (0.01, 0.88) | 0.11 (0.0, 0.35) | 0.16 (0.0, 0.88) |
| Once | slope | 0.89 (0.66, 1) | 0.86 (0.73, 1.04) | 0.85 (0.62, 1.04) | 0.79 (0.62, 0.93) | 0.76 (0.48, 0.93) |
| | $R^2$ | 0.74 (0.41, 0.97) | 0.73 (0.11, 0.96) | 0.51 (0.01, 0.93) | 0.23 (0.0, 0.67) | 0.32 (0.01, 0.92) |
| Yearly | slope | 0.97 (0.83, 1.11) | 0.94 (0.75, 1.11) | 0.97 (0.77, 1.12) | 0.93 (0.77, 1.03) | 0.92 (0.66, 1.16) |
| | $R^2$ | 0.74 (0.12, 0.98) | 0.75 (0.49, 0.95) | 0.62 (0.27, 0.97) | 0.48 (0.01, 0.82) | 0.55 (0.0, 0.91) |
| Monthly | slope | 1.05 (0.95, 1.14) | 1.03 (0.91, 1.2) | 1.03 (0.92, 1.24) | 0.97 (0.8, 1.08) | 0.96 (0.76, 1.09) |
| | $R^2$ | 0.99 (0.96, 1) | 0.99 (0.96, 1) | 0.97 (0.88, 1) | 0.89 (0.46, 0.99) | 0.85 (0.09, 0.99) |

**Figure 2.**
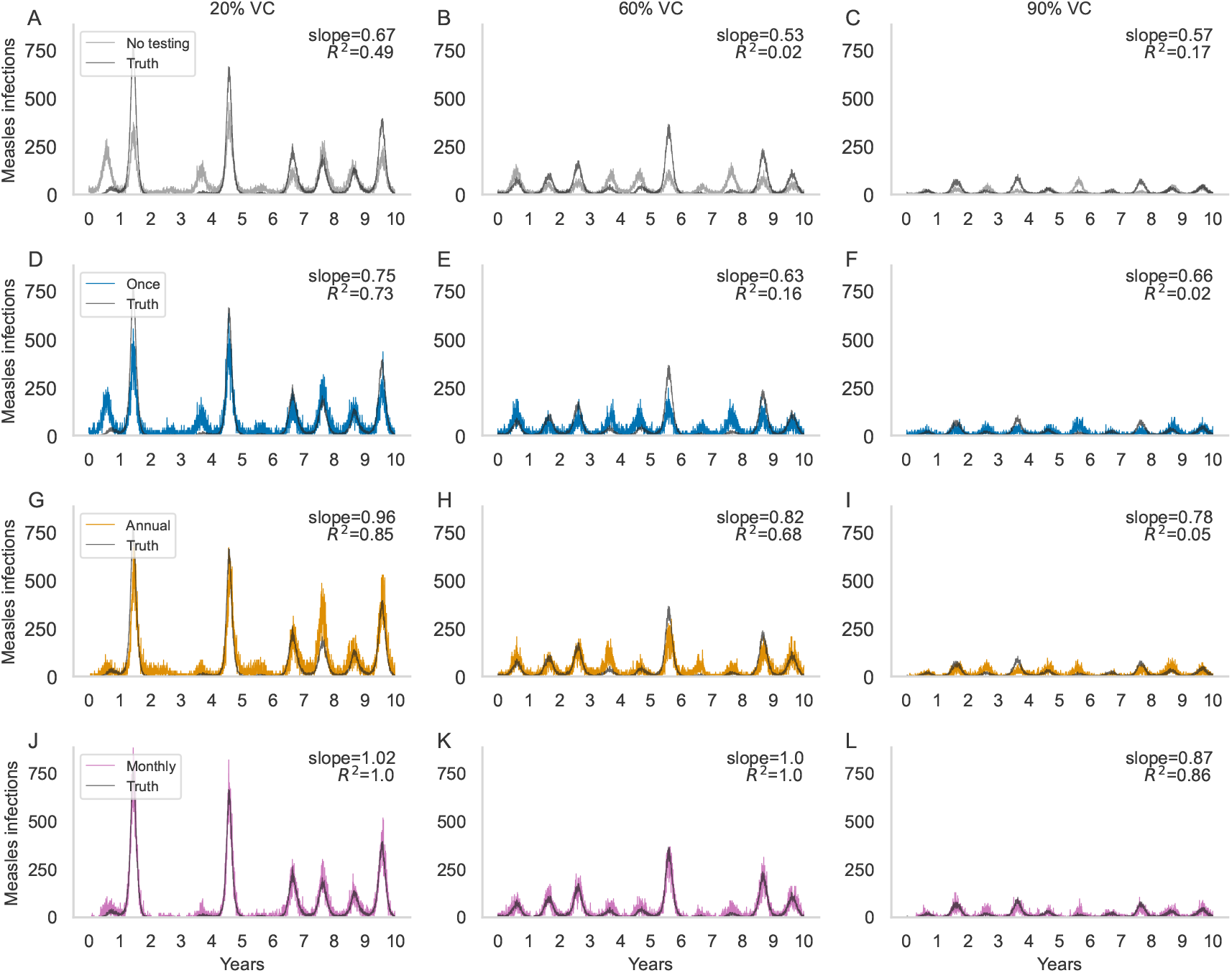
Reconstructed measles infections for different vaccination coverage (columns) by treating all febrile-rash cases as measles-positive (A–C) and by estimation with once (D–F), annual (G–I), and monthly (J–L) moving windows. Slope and *R*^2^ show the regression of the estimated annual incidence against the true incidence of the simulated time series.

The bias in the estimated reporting rate is resolved at all levels of vaccination coverage when we first reconstruct the estimated time series of measles infections using the relative proportion of cases that are measles-positive (Table 2). This holds at all testing rates (20–100% of cases tested) for a 100% sensitive and specific test and a test with 90% sensitivity and 95% specificity. The estimate of the reporting rate is robust to reconstruction of the estimated time series of measles infections based on a single estimate of the relative proportion attributable to measles for the entire time series, or rolling estimates using an annual or monthly moving window (Table 2). The estimated reporting rate has a slight positive bias (lower bound does not contain the true reporting rate of 10%) for a test with 90% sensitivity and 95% specificity when vaccination is 90%, even with 100% testing; at this level, measles prevalence is low and the positive predictive value of a diagnostic positive is lower. Note that, at 20% testing the confidence interval for the reporting rate is wider, which may contain the truth because of lower precision, despite the same bias as at 100% testing.

Though the estimate of the reporting rate is robust to the choice of time scale for estimating the relative proportion attributable to measles, the estimate of annual incidence of measles is improved using a monthly compared to annual moving window, and both perform better than a single estimate (Table 3); we present this here for a test with 90% sensitivity and 95% specificity, but the pattern is general. The slope of the regression of annual estimated measles incidence on true measles incidence decreases from 0.89 to 0.76 (i.e., the bias gets larger) as vaccination coverage increases from 20–90% when the time series is reconstructed based on a single estimate of the relative proportion (Table 3). This means that the average annual incidence of measles is increasingly under-estimated as vaccination coverage increases. The *R*^2^ decreases from 0.74 to 0.32 (Table 3). This arises because individual years with and without rubella outbreaks are treated the same with a single estimate of the proportion of cases attributable to measles. Using an annual or monthly moving window for the proportion of suspected cases attributable to measles results in lower bias in the estimate of annual incidence (slope closer to 1) and less inter-annual variability in the estimate (*R*^2^ closer to 1; Table 3). Notably, using a monthly moving window estimate of the proportion of suspected cases attributable to measles results in *R*^2^ *>* 0.85 for all levels of vaccination (Table 3). Note that as vaccination increases, the variability in measles outbreak timing and magnitude increases; thus the relative proportion of suspected cases attributable to measles becomes more variable.

Restricting only to the reconstruction of measles infections using a monthly moving window estimate, we can compare the performance of alternative test sensitivity and specificity. At low vaccination coverage (and correspondingly, higher measles prevalence) the estimator of annual incidence is comparable for test sensitivity and specificity between 85–100% (Table 4). As vaccination increases and measles prevalence declines, the estimator of annual measles incidence remains unbiased for all levels of sensitivity and specificity, as confidence intervals for the slope contains 1. However, the variation in the estimate of the slope and the inter-annual variability (lower *R*^2^) increase as test specificity decreases and vaccination increases. At lower vaccination coverage (and hence higher measles prevalence) a test with lower sensitivity and specificity provides a good estimate of the annual measles incidence. However, at higher coverage (and lower measles prevalence), the inter-annual variability when using tests with lower sensitivity and specificity declines more quickly than for high sensitivity and specificity tests. We highlight this in Table 4 by illustrating in bold, for each level of vaccination at 20% sampling, the test sensitivity and specificity below which *R*^2^ drops to 0.85 or under. For the minimal sensitivity and specificity (85% and 85% respectively), increasing the sampling rate from 20% to 60% of suspected cases does not improve the performance of the estimator. This implies that the additional noise arises because of the inclusion of false positives in the estimate of the relative proportion of suspected cases attributable to measles as relative measles prevalence declines (Table 1), rather than noise due to low sample sizes.

**Table 4:** Slope and coefficient of determination (*R*^2^) of the regression of estimated annual incidence of measles against the true simulated incidence of measles infection as a function of measles vaccination coverage (columns) and the sensitivity and specificity of tests (rows). Slope near 1 indicates low bias in estimates of annual incidence. *R*^2^ near 1 indicates low variation in estimates from the true values. Values in brackets show the range over 20 replicate time series. Bolded entries are used to guide discussion in the main text. (Monthly estimation of the proportion of cases attributable to measles)

| Monthly scenario |  |  | Vaccination Coverage |  |  |  |  |
| --- | --- | --- | --- | --- | --- | --- | --- |
| Sensitivity/specificity | Sampling |  | 20% | 40% | 60% | 80% | 90% |
| 100/100 | 20% | slope | 1.07 (0.98, 1.18) | 1.06 (0.94, 1.25) | 1.07 (0.97, 1.25) | 1.03 (0.85, 1.16) | **1.04 (0.91, 1.18)** |
| | | $R^2$ | 0.99 (0.92, 1) | 0.99 (0.97, 1) | 0.98 (0.95, 1) | 0.96 (0.81, 0.99) | **0.97 (0.85, 1)** |
| 90/95 | 20% | slope | 1.05 (0.95, 1.14) | 1.03 (0.91, 1.2) | 1.03 (0.92, 1.24) | **0.97 (0.8, 1.08)** | 0.96 (0.76, 1.09) |
| | | $R^2$ | 0.99 (0.96, 1) | 0.99 (0.96, 1) | 0.97 (0.88, 1) | **0.89 (0.46, 0.99)** | 0.85 (0.09, 0.99) |
| 90/90 | 20% | slope | 1.04 (0.92, 1.15) | 1.04 (0.87, 1.18) | 1.0 (0.89, 1.17) | 0.95 (0.8, 1.07) | 0.91 (0.66, 1.03) |
| | | $R^2$ | 0.99 (0.96, 1) | 0.98 (0.83, 1) | 0.94 (0.77, 1) | 0.8 (0.14, 0.96) | 0.74 (0.01, 0.99) |
| 85/90 | 20% | slope | 1.03 (0.93, 1.15) | 1.03 (0.86, 1.19) | **1.0 (0.89, 1.18)** | 0.94 (0.79, 1.07) | 0.91 (0.7, 1.05) |
| | | $R^2$ | 0.98 (0.93, 1) | 0.98 (0.83, 1) | **0.93 (0.75, 1)** | 0.78 (0.01, 0.97) | 0.72 (0.01, 0.98) |
| 90/85 | 20% | slope | 1.01 (0.87, 1.11) | 1.0 (0.82, 1.16) | 0.97 (0.86, 1.12) | 0.93 (0.77, 1.07) | 0.88 (0.61, 1.01) |
| | | $R^2$ | 0.97 (0.91, 1) | 0.95 (0.75, 1) | 0.85 (0.2, 0.99) | 0.69 (0.0, 0.95) | 0.64 (0.0, 0.98) |
| 85/85 | 20% | slope | **1.0 (0.89, 1.1)** | **0.98 (0.86, 1.17)** | 0.98 (0.85, 1.15) | 0.91 (0.73, 1.03) | 0.87 (0.63, 1.04) |
| | | $R^2$ | **0.96 (0.85, 1)** | **0.95 (0.79, 0.99)** | 0.84 (0.32, 0.99) | 0.67 (0.0, 0.92) | 0.6 (0.06, 0.96) |
| 85/85 | 40% | slope | 1.01 (0.88, 1.13) | 0.99 (0.83, 1.15) | 0.97 (0.84, 1.11) | 0.91 (0.78, 1.05) | 0.87 (0.6, 1.03) |
| | | $R^2$ | 0.98 (0.89, 1) | 0.95 (0.66, 1) | 0.86 (0.29, 1) | 0.67 (0.02, 0.96) | 0.63 (0.0, 0.97) |
| 85/85 | 60% | slope | 1.0 (0.87, 1.11) | 0.99 (0.84, 1.14) | 0.97 (0.83, 1.11) | 0.91 (0.77, 1.05) | 0.87 (0.59, 1) |
| | | $R^2$ | 0.98 (0.89, 1) | 0.95 (0.67, 1) | 0.87 (0.48, 1) | 0.69 (0.02, 0.96) | 0.63 (0.0, 0.98) |

These patterns are robust to a higher birth rate (Table S4); lower case importation rates (Table S2), which increases variability; and assuming assortative mixing (Table S3), which is inconsistent with the implicit assumption of the TSIR model, reduces the performance of the estimator overall, but nonetheless does not interact with the test characteristics.

## 4 Discussion

The decreasing specificity of the measles clinical case definition with increasing vaccination coverage means that, in the absence of diagnostic confirmation, clinical febrile rash surveillance will dramatically over-estimate the reporting rate and under-estimate the burden of measles disease. The magnitude of this error increases as vaccination coverage increases and measles becomes a decreasing fraction of all clinical febrile rash. Here we show that relatively limited rates of diagnostic confirmation applied systematically can overcome this bias and produce reliable estimates of total measles burden by using the proportion of tested cases that are confirmed as measles-positive to estimate the proportion of untested cases that are attributable to measles.

As measles vaccination coverage increases, so too does the inter-annual variability of measles incidence (as described in [9]). Thus, even at constant levels of vaccination, the relative proportion of clinically compatible cases that are true measles cases is expected to vary significantly over time. In our simulations, estimating the proportion of suspected measles cases that are attributable to measles using recently tested cases (e.g. on a 1-month moving window) resulted in more accurate estimates (low bias, low variance) of annual measles incidence.

The sensitivity and specificity of the diagnostic test used affects the performance of the estimator of measles incidence. This follows the well known relationship between the positive predictive value of a diagnostic test, the specificity of the test, and the prevalence of the outcome. When vaccination coverage is low and measles is at high prevalence there is little difference in the accuracy of estimates of the annual incidence of measles using tests that span the likely range of diagnostic performance for measles IgM RDTs and EIAs (sensitivity and specificity *>* 85%). However, as vaccination coverage increases and measles decreases in prevalence relative to other sources of suspected cases, the proportion of tested cases that are positive becomes an increasingly biased estimator of the true prevalence of measles among suspected cases, and that bias is larger for tests with lower accuracy. The bias increases with decreasing sensitivity and specificity, but is more impacted by the latter. This implies that, as vaccination coverage increases, and measles becomes rarer, there should be an emphasis on using diagnostics with higher accuracy. However, where and when to switch between lower accuracy and higher accuracy tests will depend on operational considerations such as cost and availability.

We note that, in the case of measles, the introduction of rubella-containing vaccine as measles vaccination coverage increases means that positive predictive value of any diagnostic test should increase (because the pre-test odds of a suspected case being rubella should decrease) and the performance of lower accuracy diagnostics will be expected to improve relative to the scenario without rubella vaccine.

Diagnostic uncertainty presents a challenge to the estimation of disease burden for infections, like measles, that present with common symptoms. However, increased access to diagnostic confirmation, even with imperfect assays, can help to produce reliable estimates of overall disease burden. Though we have presented this case study in the context of measles and febrile rash surveillance, the general patterns would be applicable to any non-specific syndromic surveillance system.

## Data Availability

All data produced in the present study are available upon reasonable request to the authors.

## Acknowledgements

We acknowledge grant funding from Gavi, the Vaccine Alliance.

## Author contributions

M.F. conceptualized, supervised, and acquired funding for the study. T.L. developed the model, analyzed the data, and wrote the first draft of the manuscript. M.F. and T.L. contributed to the final draft.

## Supplementary Information for: Combining clinical and diagnostic surveillance to estimate the burden of measles disease: a modeling study

Tiffany Leung, Matthew Ferrari

### Mathematical model of measles and rubella

We constructed an age-structured stochastic compartmental model of measles and rubella transmission with vaccination (Figure S1). The population is divided into 40 age groups (24 monthly age groups from 0 to 2 years old; 1 age group from 2 to 5 years old; 14 five-year age groups from 5 to 75 years old; and 1 age group for 75 years and older). Individuals are either unvaccinated *U*_*mr*_ or vaccinated, *V*_*mr*_, where *m* and *r* further describe the immune status for measles and for rubella respectively. Individuals transition through the immune status compartments for measles (*m*) and for rubella (*r*): temporarily immune with maternal antibodies (M), susceptible (S), infectious (I), or recovered and immune (R). For example, an individual *U*_*SI*_ represents someone who is unvaccinated, susceptible to measles and infected with rubella.

Individuals are born either susceptible or temporarily immune with maternally antibodies to measles and/or to rubella, as governed by the birth rate and the proportional population-level immunity of the fertile age groups between 10 and 55 years old [13]. All individuals face a constant death rate. Individuals age exponentially through discrete age classes. For each disease, we assumed lifelong immunity following infection.

The probability of vaccination is calculated as a cumulative probability that an individual would be vaccinated by a specific age. The probability is then scaled to the vaccination coverage at the specific age. We use a one-dose vaccine with 94% vaccine efficacy against measles infection and ineffective against rubella. We used a truncated normally distributed with a mean of 9 months and standard deviation of 0.5 months. The distribution is truncated at 0 and 24 months.

The model for the first age class of the unvaccinated population (with births into the population) can be described by a set of ordinary differential equations:

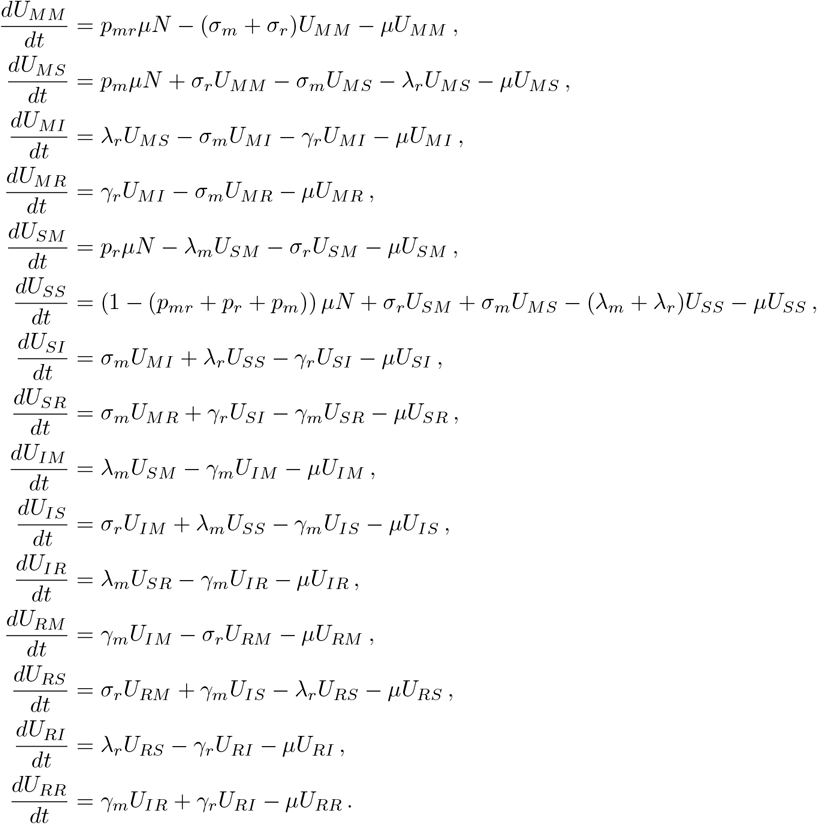

The equations of the force of infection for measles and for rubella for age class *i* at time *t* are, respectively,

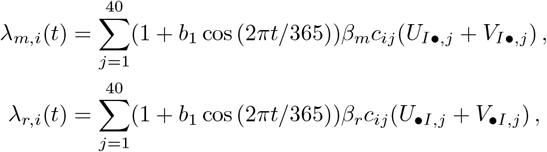

where *b*_1_ is the relative amplitude of seasonal forcing, *β*_*m*_, *β*_*r*_ are the transmission coefficients for measles and rubella, and *c*_*ij*_ is the contact rate from age group *j* to age group *i*. Births enter the population through four unvaccinated states:

- *U*_*MM*_, maternally immune to measles and rubella;
- *U*_*MS*_, maternally immune to measles and susceptible to rubella;
- *U*_*SM*_, susceptible to measles and maternally immune to rubella;
- *U*_*SS*_, susceptible to measles and rubella

as governed by the proportion, *p* of the population of fertile age that is immune (i.e., mothers pass immunity to the child). Equations for the vaccinated classes are the same as ones for the unvaccinated but without births. A table of parameters is presented in Table S1.

## Tables

Here we present the slope and *R*^2^ of the regression of the estimated annual incidence of measles against the true simulated incidence of measles infection for various scenarios. Here we use 20% testing rate and a test with 90% sensitivity and 95% specificity.

### Lower case importation

Table S2 shows the regression of the estimated annual incidence of measles against the true simulated incidence of measles infection when lower case importation is 0.0015 and 0.00075, equivalent to a 50% and 75% reduction of the default case importation rate respectively.

### Assortative mixing

Table S3 shows the regression of the estimated annual incidence of measles against the true simulated incidence of measles infection under Kenya-like assortative contact mixing [15].

### Higher birth rate

Table S4 shows the regression of the estimated annual incidence of measles against the true simulated incidence of measles infection under an annual birth rate of 40 per 1,000 persons.

**Figure S1:**
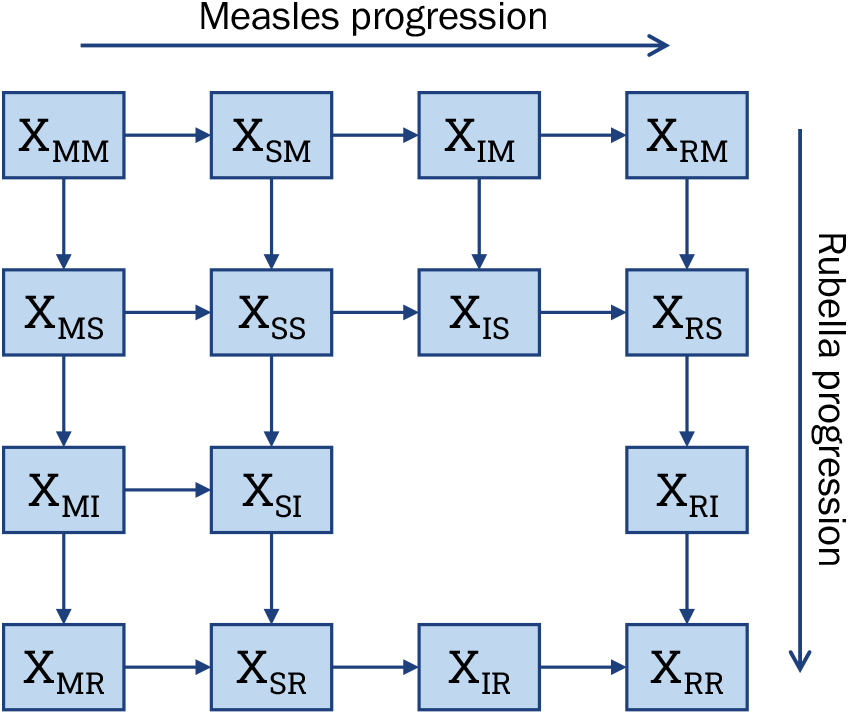
Model diagram of measles and rubella transmission. Individuals are grouped into classes *X*_*m*,*r*_, where *X* is their vaccination status (unvaccinated or vaccinated; *X* = *U, V*). The vaccination status is further divided by immune status to measles (*m*) and to rubella (*r*), which are: temporarily immune with maternal antibodies (M), susceptible (S), infectious (I) and recovered (R).

**Table S1:**
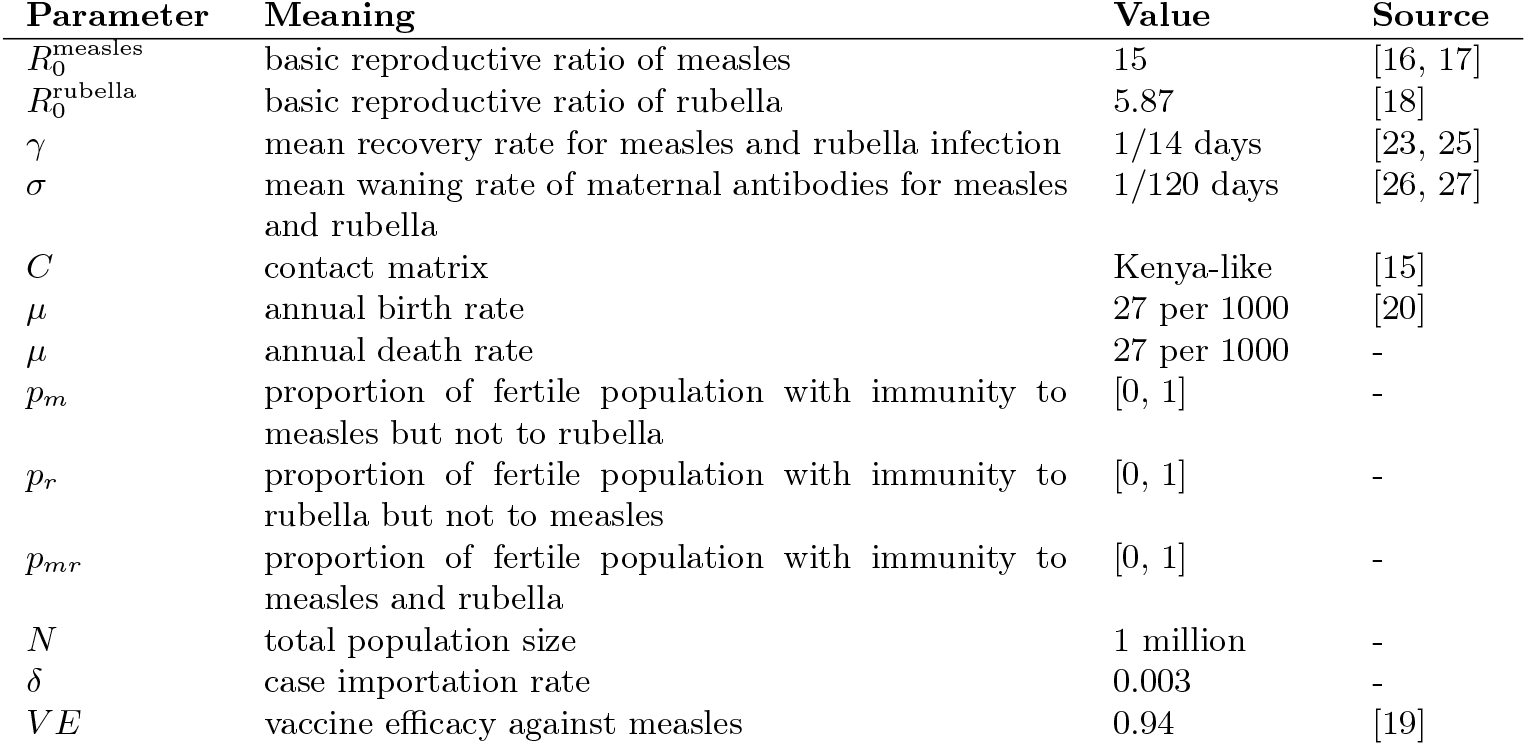
Description of model parameters.

| Parameter | Meaning | Value | Source |
| --- | --- | --- | --- |
| $R_0^{\text{measles}}$ | basic reproductive ratio of measles | 15 | [16, 17] |
| $R_0^{\text{rubella}}$ | basic reproductive ratio of rubella | 5.87 | [18] |
| $\gamma$ | mean recovery rate for measles and rubella infection | 1/14 days | [23, 25] |
| $\sigma$ | mean waning rate of maternal antibodies for measles and rubella | 1/120 days | [26, 27] |
| $C$ | contact matrix | Kenya-like | [15] |
| $\mu$ | annual birth rate | 27 per 1000 | [20] |
| $\mu$ | annual death rate | 27 per 1000 | - |
| $p_m$ | proportion of fertile population with immunity to measles but not to rubella | [0, 1] | - |
| $p_r$ | proportion of fertile population with immunity to rubella but not to measles | [0, 1] | - |
| $p_{mr}$ | proportion of fertile population with immunity to measles and rubella | [0, 1] | - |
| $N$ | total population size | 1 million | - |
| $\delta$ | case importation rate | 0.003 | - |
| $VE$ | vaccine efficacy against measles | 0.94 | [19] |

**Table S2:** Slope and coefficient of determination (*R*^2^) of the regression of estimated annual incidence of measles against the true simulated incidence of measles infection as a function of measles vaccination coverage (columns) and the time scale for estimating the proportion of cases attributable to measles (rows) at lower case importation rates. Slope near 1 indicates low bias in estimates of annual incidence. *R*^2^ near 1 indicates low variation in estimates from the true values. Values in brackets show the range over 20 replicate time series.

| Case importation: 0.0015 |  | Vaccination Coverage |  |  |  |  |
| --- | --- | --- | --- | --- | --- | --- |
|  |  | 20% | 40% | 60% | 80% | 90% |
| All | Slope | 0.84 (0.63, 1.01) | 0.79 (0.55, 0.96) | 0.73 (0.51, 1.04) | 0.64 (0.39, 0.89) | 0.67 (0.28, 0.94) |
| | $R^2$ | 0.51 (0.01, 0.94) | 0.42 (0.0, 0.81) | 0.23 (0.0, 0.86) | 0.17 (0.0, 0.83) | 0.17 (0.0, 0.76) |
| Once | Slope | 0.89 (0.7, 1.03) | 0.85 (0.64, 1.01) | 0.8 (0.58, 1.08) | 0.72 (0.48, 0.94) | 0.75 (0.38, 1) |
| | $R^2$ | 0.66 (0.11, 0.98) | 0.59 (0.04, 0.92) | 0.45 (0.01, 0.93) | 0.3 (0.0, 0.92) | 0.28 (0.0, 0.82) |
| Yearly | Slope | 0.97 (0.77, 1.14) | 0.96 (0.83, 1.1) | 0.93 (0.75, 1.14) | 0.9 (0.7, 1.08) | 0.94 (0.6, 1.23) |
| | $R^2$ | 0.74 (0.5, 0.93) | 0.72 (0.08, 0.95) | 0.6 (0.07, 0.94) | 0.56 (0.05, 0.91) | 0.57 (0.2, 0.9) |
| Monthly | Slope | 1.06 (0.92, 1.22) | 1.06 (0.95, 1.17) | 1.02 (0.83, 1.27) | 0.94 (0.77, 1.09) | 0.97 (0.73, 1.19) |
| | $R^2$ | 0.99 (0.97, 1) | 0.98 (0.93, 1) | 0.96 (0.74, 1) | 0.91 (0.6, 0.99) | 0.88 (0.66, 0.99) |
| Case importation: 0.00075 |  |  |  |  |  |  |
| All | Slope | 0.75 (0.47, 1.01) | 0.73 (0.46, 1.03) | 0.7 (0.43, 1.07) | 0.61 (0.4, 0.88) | 0.54 (0.2, 0.83) |
| | $R^2$ | 0.43 (0.0, 0.94) | 0.34 (0.04, 0.87) | 0.37 (0.01, 0.81) | 0.27 (0.0, 0.79) | 0.25 (0.02, 0.83) |
| Once | Slope | 0.82 (0.57, 1.02) | 0.81 (0.6, 1.06) | 0.77 (0.54, 1.11) | 0.69 (0.5, 0.91) | 0.63 (0.31, 0.89) |
| | $R^2$ | 0.62 (0.21, 0.98) | 0.54 (0.0, 0.95) | 0.54 (0.01, 0.93) | 0.43 (0.0, 0.89) | 0.39 (0.0, 0.92) |
| Yearly | Slope | 0.95 (0.76, 1.2) | 0.98 (0.77, 1.22) | 0.92 (0.71, 1.13) | 0.89 (0.7, 1.17) | 0.84 (0.52, 1.08) |
| | $R^2$ | 0.81 (0.6, 0.96) | 0.77 (0.01, 0.99) | 0.7 (0.02, 0.92) | 0.71 (0.29, 0.98) | 0.67 (0.0, 0.95) |
| Monthly | Slope | 1.06 (0.86, 1.36) | 1.08 (0.93, 1.35) | 1.01 (0.85, 1.28) | 0.95 (0.79, 1.12) | 0.91 (0.65, 1.19) |
| | $R^2$ | 0.99 (0.97, 1) | 0.98 (0.95, 1) | 0.97 (0.83, 1) | 0.96 (0.8, 0.99) | 0.91 (0.49, 1) |

**Table S3:** Slope and coefficient of determination (*R*^2^) of the regression of estimated annual incidence of measles against the true simulated incidence of measles infection as a function of measles vaccination coverage (columns) and the time scale for estimating the proportion of cases attributable to measles (rows) with assortative mixing. Slope near 1 indicates low bias in estimates of annual incidence. *R*^2^ near 1 indicates low variation in estimates from the true values. Values in brackets show the range over 20 replicate time series.

| Assortative mixing |  | Vaccination Coverage |  |  |  |  |
| --- | --- | --- | --- | --- | --- | --- |
|  |  | 20% | 40% | 60% | 80% | 90% |
| All | Slope | 0.79 (0.68, 1.03) | 0.67 (0.51, 0.8) | 0.62 (0.49, 0.79) | 0.54 (0.36, 0.65) | 0.5 (0.29, 0.68) |
| | $R^2$ | 0.54 (0.06, 0.98) | 0.34 (0.0, 0.94) | 0.1 (0.0, 0.51) | 0.13 (0.0, 0.55) | 0.24 (0.0, 0.79) |
| Once | Slope | 0.88 (0.78, 1.07) | 0.77 (0.62, 0.87) | 0.75 (0.64, 0.89) | 0.66 (0.49, 0.77) | 0.61 (0.42, 0.78) |
| | $R^2$ | 0.72 (0.13, 0.99) | 0.57 (0.0, 0.98) | 0.28 (0.0, 0.77) | 0.15 (0.0, 0.66) | 0.27 (0.0, 0.84) |
| Yearly | Slope | 0.92 (0.79, 1.07) | 0.83 (0.71, 1.05) | 0.84 (0.7, 0.99) | 0.79 (0.63, 0.9) | 0.76 (0.59, 0.91) |
| | $R^2$ | 0.62 (0.01, 0.93) | 0.6 (0.01, 0.96) | 0.38 (0.0, 0.88) | 0.21 (0.0, 0.85) | 0.3 (0.0, 0.87) |
| Monthly | Slope | 1.07 (1.03, 1.1) | 0.95 (0.9, 0.99) | 0.95 (0.88, 1.03) | 0.86 (0.79, 0.95) | 0.82 (0.76, 0.95) |
| | $R^2$ | 0.95 (0.45, 1) | 0.96 (0.72, 1) | 0.96 (0.84, 0.99) | 0.82 (0.11, 0.98) | 0.81 (0.48, 0.98) |

**Table S4:** Slope and coefficient of determination (*R*^2^) of the regression of estimated annual incidence of measles against the true simulated incidence of measles infection as a function of measles vaccination coverage (columns) and the time scale for estimating the proportion of cases attributable to measles (rows) with a higher birth rate of 40 births per 1000 persons. Slope near 1 indicates low bias in estimates of annual incidence. *R*^2^ near 1 indicates low variation in estimates from the true values. Values in brackets show the range over 20 replicate time series.

| 40 births per 1000 |  | Vaccination Coverage |  |  |  |  |
| --- | --- | --- | --- | --- | --- | --- |
|  |  | 20% | 40% | 60% | 80% | 90% |
| All | Slope | 0.85 (0.62, 0.97) | 0.79 (0.63, 0.94) | 0.67 (0.46, 0.86) | 0.71 (0.55, 0.83) | 0.63 (0.45, 0.85) |
| | $R^2$ | 0.37 (0.01, 0.92) | 0.3 (0.0, 0.78) | 0.27 (0.0, 0.9) | 0.26 (0.0, 0.7) | 0.11 (0.0, 0.44) |
| Once | Slope | 0.9 (0.7, 1.04) | 0.86 (0.72, 1.02) | 0.77 (0.59, 0.97) | 0.8 (0.67, 0.93) | 0.72 (0.57, 0.97) |
| | $R^2$ | 0.58 (0.08, 0.97) | 0.51 (0.09, 0.91) | 0.43 (0.03, 0.97) | 0.33 (0.0, 0.77) | 0.14 (0.0, 0.56) |
| Yearly | Slope | 0.94 (0.78, 1.11) | 0.93 (0.74, 1.07) | 0.9 (0.76, 1.14) | 0.95 (0.77, 1.1) | 0.88 (0.76, 1.15) |
| | $R^2$ | 0.61 (0.02, 0.91) | 0.53 (0.04, 0.92) | 0.59 (0.04, 0.93) | 0.43 (0.0, 0.93) | 0.33 (0.01, 0.91) |
| Monthly | Slope | 1.04 (0.95, 1.22) | 1.04 (0.98, 1.19) | 1.01 (0.91, 1.22) | 0.97 (0.91, 1.08) | 0.93 (0.83, 1.22) |
| | $R^2$ | 0.98 (0.94, 1) | 0.98 (0.93, 1) | 0.97 (0.93, 1) | 0.86 (0.45, 0.99) | 0.81 (0.54, 0.98) |

